# Prevalence and Risk factors of Urinary Schistosomiasis in Kaporo Village, Karonga District, Malawi

**DOI:** 10.1101/2023.06.05.23290821

**Authors:** Christopher S. Nyondo, Rhosheen Mthawanji, Master Chisale

## Abstract

**Objective:** The aim of the study was to determine the prevalence and Risk factors of urogenital schistosomiasis in Karonga district; it also sought to determine the relationship between involvement in MDA advocacy campaigns and Knowledge level of the disease.

**Methods:** The study enrolled 251 participants that responded to the questionnaire-guided interview and submitted urine for microscopy.

**Results:** Of 251 children that were enrolled 87 (34.7%) were found to have *S.haematobium* eggs. Chi-square analysis established that having a parent in rice farming (p=0.029) occupation is a key risk factor for urogenital schistosomiasis. It was also surprising to note that those schoolchildren who received Praziquantel during MDA had significantly higher prevalence (p=0.010). Furthermore, this study revealed that they is no association between a child involving in MDA advocacy compaigns and level of knowledge on schistosomiasis transmission.

## Introduction

Schistosomiasis is a water-borne chronic parasitic disease caused by trematode worms of the genus Schistosoma(1). Despite Malawi government introducing mass drug administration in schools as a measure of controlling the infection, the prevalence rate of the infection in children is still high (2). Among neglected parasitic diseases, schistosomiasis remains the most prevalent in the world and is reported endemic in almost 76 countries (3). *Schistosoma haematobium* and *Schistosoma mansoni* are the predominant species in Africa, causing urogenital and intestinal schistosomiasis respectively. Humans get infection while engaging in recreation, domestic and occupation activities. Among the high risk group includes; pre-school, school going children, irrigation workers and fishermen. Also known to be more prevalent on those who live in poor conditions particularly in terms of water and sanitation (4).World Health Organization (WHO) recommends that young children in endemic areas be treated with praziquantel and therefore introduced mass drug administration (MDA) with praziquantel as intervention for schistosomiasis control in many endemic countries including Malawi(5). Reports shows that with MDA, prevalence have drastically been reduced in the past few decades. However despite MDA interventions, high prevalence has been reported in some parts of Southern Malawi(3). This only shows that ever since Malawi introduced MDA in endemic areas, schistosomiasis prevalence post MDA era remains unclear hence the urgent need of epidemiological data on the current prevalence in endemic districts of Malawi.

Most of the published schistosomiasis epidemiological studies have been done in southern and Eastern region of Malawi, where high prevalence has been reported. This has an impact on the national evaluation of the schistosomiasis control interventions such as MDA. Malawi is blessed with several water bodies which scholars estimated that at least 20% are infected with schistosomiasis(5). Malawi ministry of health set targets to eliminate the disease by 2020, however the target hit the dead rock as high prevalence have been reported in other areas posts the set targeted year. Now that, World Health Organization in its 2030 strategic plan aims to eliminate most NTDs including schistosomiasis in most endemic countries, certainly Malawi will adopt the plans and that’s therefore need to step up and provide up to date information on the epidemiology of the disease in both vector and human hosts.

In order to estimate the burden of the disease, and asses the possible impact of MDA, it remains important to collect up to date information to assist in planning. Therefore, the study aimed at providing prevalence estimates for school going children in Karonga and assessing the possible risk factors including establishing the relationship between involvement in MDA advocacy campaigns with knowledge on urinary schistosomiasis.

## Main text

### Study area and period

This study took place around Kaporo rural hospital catchment area in Karonga district in northern Malawi, from December 2021 to January 2023. Kaporo area is a flat area blessed with rivers and lakes that usually floods during rainy season. The main activities of People in the area are rice farming and fishing.

### Type of research study

A cross-sectional study that involved both Questionnaire guided interview among school going children and laboratory urine analysis for schistosomiasis eggs identification was employed.

### Study population

The target population of this study were school going children aged ≤15 years in areas around Kaporo rural hospital catchment area and have lived in the area for at least one year prior to the study.

### Sample size and sampling techniques

Schistosomiasis prevalence was determined, with a precision of 7% and a 95% Confidence Interval and the prevalence rate previously registered in the same area of 21% (NTD master plan, 2015-2020) was used for a 10,000 finite primary school children. Therefore, using online-based software, the estimated sample size was 196 or more pupils. This study therefore, enrolled 272 participants from 3 schools randomly selected, of that 13 (4.8%) participants retained empty bottles and 8 (2.9%) participants denied in participating. Therefore, 251 participants (92.3%) completed the study and were considered in analysis.

### Data collection and Laboratory examination

A questionnaire-guided interview was used to collect each child’s demographics, water contact behavior, knowledge on schistosomiasis, parent’s education and occupation including history of receiving anti-schistosoma treatment. Knowledge on the disease was assessed by interviewing the children on the modes of transmission (contact with unsafe water sources potentially infested with cercariae), symptoms of the disease (blood in urine, painful urination and supra pubic pain), availability of treatment (can be treated by modern medicine) and control measures (avoid urination in water sources, avoid contact with unsafe water sources, use of safe source of water for domestic use from taps and wells, participation in MDA advocacy campaigns). The questionnaire was coded in a software called Kobo data collector and pre-tested for all skip logic and efficiency in exporting to SPSS or Excel. A clean wide mouthed container was given to each child in which terminal voided urine was provided around 10am – 2pm. collected samples was then immediately examined at Karonga district hospital using sedimentation techniques (13). The results were shared with Karonga district Hospital and Kaporo rural hospital, another round of Praziquantel administration was done in the participated schools following the study outcome.

### Data analysis

Data was analyzed using IBM SPSS statistic 20. Presence of schistosomiasis, demographic, water contact, parent’s education and occupation were treated as categorical variables and presented as frequencies and percentages. Knowledge was assessed on a scale of 9 points score with categories: 1-3 considered as low, 4-6 medium and 7-9 as high knowledge. The categories followed correct mentioning of the modes of transmission (contact with cercariae infested waters), symptoms of the disease (blood in urine, painful urination and suprapubic pain), availability of treatment (can be treated by modern medicine) and control measures (avoid urination in water sources, avoid contact with unsafe water sources, use of safe source of water such as taps and wells for domestic use, participation in MDA advocacy campaigns) to constitute 9 points score. For inferential statistics, the dependent variable was schistosomiasis whereas the independent variables were the demographic factors (age and gender), socioeconomic factors (fathers’ educational levels, parents ‘employment status). Chi-square test was used to examine the significance of the association. A p-value of ≤ 0.05 was considered significant. Factors that came out significant were further analyzed using ordinal regression analysis to establish if they are risk factors.

## Results

### Demographic characteristics of the study participants

Two hundred and Fifty one school going children aged ≤ 15 years with a mean age of 11 years participated voluntarily in this study. The general characteristics of the participants and their families are shown in Table 1. Overall, more than half (78.2%) of the parents/guardians of the participants went up to at least primary level and at least half of the fathers (50.3%) are farmers. Of interesting a high percentage of mothers (61.5%) were involved in farming more than fathers; however, a good percentage (37.4%) of the mothers were not working (Housewives).

**Table 1.**
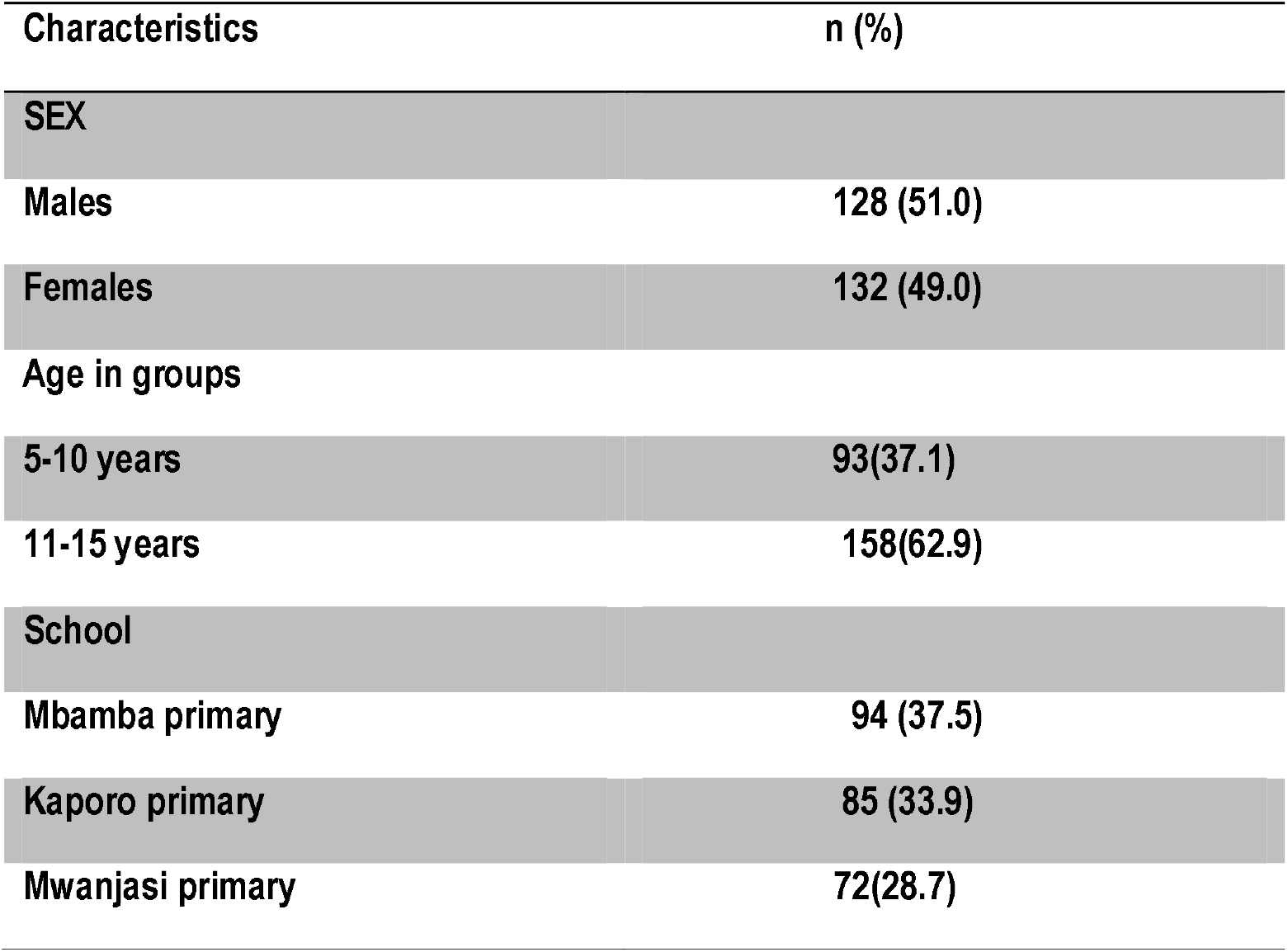
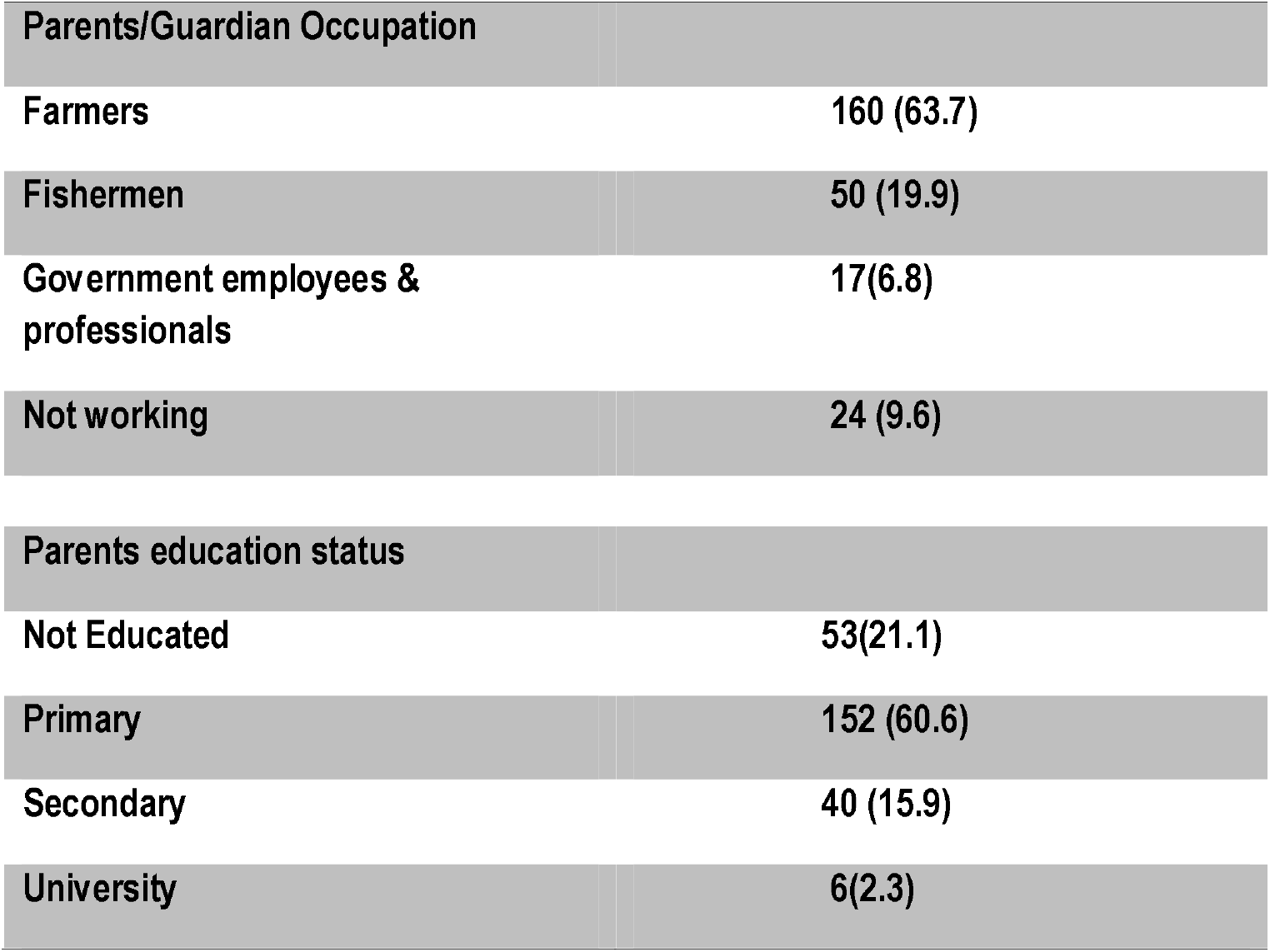
General characteristics of School going children who participated in this study (n = 251).

### Prevalence of urogenital schistosomiasis among children around Kaporo rural catchment area

Urine samples were collected from 251 participants and examined for the presence of *schistosoma haematobium* parasite ova. A total of 87(34.7%) of the participants were found to be infected with at least an egg of the parasite (fig 1). Majority (49.4%) of the children found to be infected with *S.haematobium* came from Mbamba primary school, seconded by Kaporo primary (41.4%) and a prevalence of 11.1% from Mwanjasi primary (p=0.000). Of those infected, 67 (84.8%) had light infection and 20 (15.2%) had heavy infection (p =0.00), table 2 shows the results.

**Table 2:** Prevalence and Intensity of *S. haematobium* infection by School (n=251)

| School | Number | S.haematobium present |  | Intensity |  |
| --- | --- | --- | --- | --- | --- |
|  |  | Yes | No | Light | Heavy |
| Mbamba | 94 | 43 (45.7%) | 51 (54.3%) | 35 (37.2%) | 8 (8.5%) |
| Kaporo | 85 | 36 (42.4%) | 49 (57.6%) | 32(37.6%) | 4 (4.7%) |
| Mwanjasi | 72 | 8(11.1%) | 64(88.9%) | 8 (11.1%) | 0 (0.0%) |

X<sup>2</sup> Test (schools vs. intensity) = 26.733<sup>a</sup>, p-value =0.000; X<sup>2</sup>Test (schools) =24.951, p=0.000

**Fig. 1.**
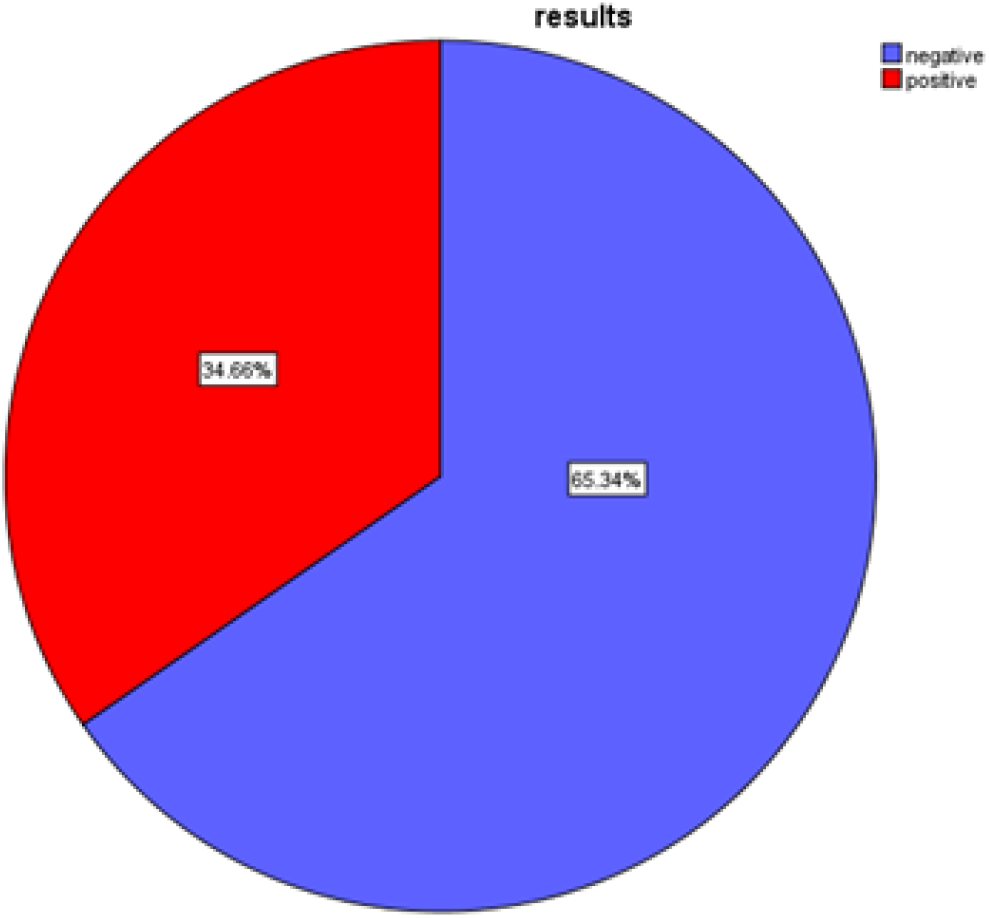
prevalence of urogenital schistosomiasis in areas around Kaporo Rural hospital

### Risk factors associated with urogenital schistosomiasis among schoolchildren in areas surrounding Kaporo rural hospital catchment area

The results for chi-square analysis for the association of presence of urogenital schistosomiasis and demographics, water contact behaviors and socioeconomic factors are shown in table 3. From the table it shows that, children whose father’s occupation is farming had significantly higher prevalence (50.0%, p-value= 0.029) than other occupations. Surprisingly, children who received praziquantel drug through mass drug administration had significantly higher prevalence (47.0%, p-value=0.010) when compared to those that did not receive the drug.

**Table 3.**
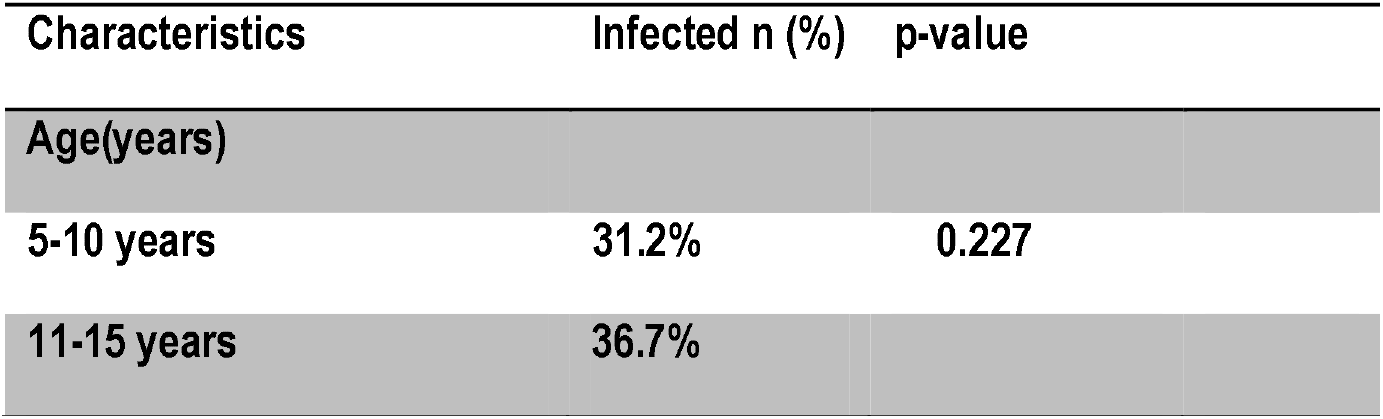
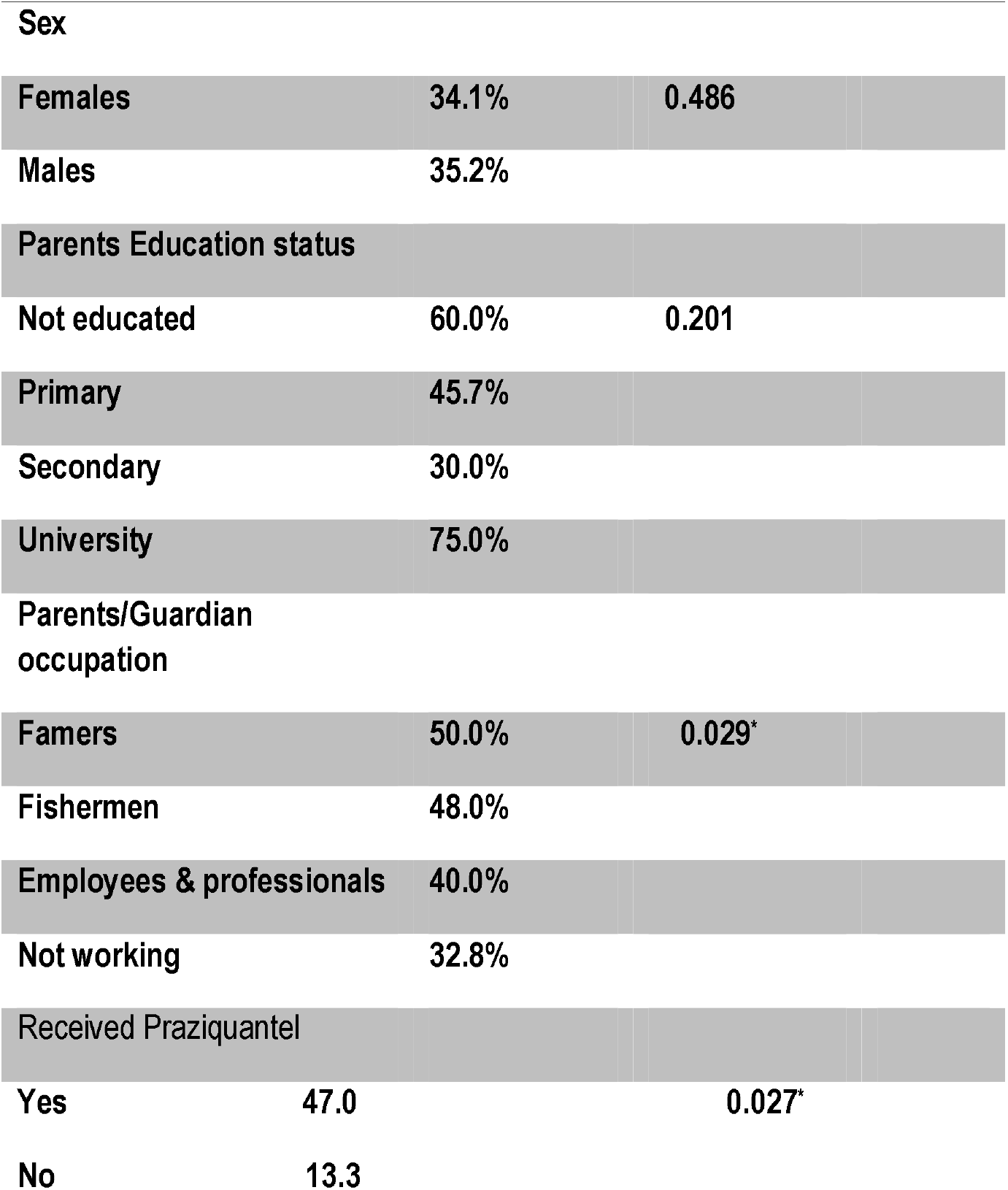
Chi-square analysis of factors associated with urogenital schistosomiasis among Kaporo rural hospital catchment area children who participated in this study (n = 179).

| Characteristics | Infected n (%) | p-value |
| --- | --- | --- |
| Age(years) |  |  |
| 5-10 years | 31.2% | 0.227 |
| 11-15 years | 36.7% |  |
| **Sex** |  |  |
| **Females** | **34.1%** | **0.486** |
| **Males** | **35.2%** |  |
| **Parents Education status** |  |  |
| **Not educated** | **60.0%** | **0.201** |
| **Primary** | **45.7%** |  |
| **Secondary** | **30.0%** |  |
| **University** | **75.0%** |  |
| **Parents/Guardian occupation** |  |  |
| **Famers** | **50.0%** | **0.029\*** |
| **Fishermen** | **48.0%** |  |
| **Employees & professionals** | **40.0%** |  |
| **Not working** | **32.8%** |  |
| **Received Praziquantel** |  |  |
| **Yes** | **47.0** | **0.027\*** |
| **No** | **13.3** |  |

### Relationship between involvement in MDA advocacy campaign and knowledge of the disease among children in Areas around Kaporo Rural hospital

Knowledge about the disease was categorized into 3 levels on a scale of 9 points score with categories: 1-3 considered as low, 4-6 medium and 7-9 as high knowledge as described in the Methodology. A high percentage (76.6%) of the schoolchildren had Low knowledge about urinary schistosomiasis (Table 4). Involvement in the MDA advocacy campaigns was not significantly associated to high knowledge about urinary schistosomiasis (p=0.190).

**Table 4.** Relationship between involvement in MDA advocacy campaigns with knowledge on urinary schistosomiasis among schoolchildren in areas around Kaporo rural hospital catchment area.

| **Involvement in MDA advocacy campaigns** | **Level of knowledge** |  |  |
| --- | --- | --- | --- |
|  | **Low** | **Medium** | **High** |
| **Yes** | **37(28.2%)** | **85(64.9%)** | **9(6.9%)** |
| No | 36(76.6%) | 7(14.9%) | 4(8.5%) |
| Total | 73(46.8%) | 92(38.0%) | 13(15.2%) |

$\chi^2$ Test= 3.319, p value = 0.190

## Discussion

Malawi national schistosomiasis prevalence is estimated to be 50% which was determined some 20 years ago (11). Many chemotherapeutic agents including introduction of mass treatment in schools and high-risk communities have been taking place since 2008 with a dream of eliminating the disease. Despite the success story of treatment of asymptomatic individuals, majority in the poor communities expose themselves to risky behaviors that put them on high chance of re-infection. Demographic characteristics (Table 1) clearly show that majority of the community members are either famers or fishermen. In a rural set up, farming activities attract everyone including the children to assist in gardens and because fishing is a financial generating activity, every male child when growing up aims for the activity hence they start practicing at very early age. The behavior increases chances of being re-infected despite having being successfully dewormed.

The present study found an overall of 34.7% against a national prevalence of 50%. This is an alarming very high prevalence, which therefore imply that urogenital schistosomiasis is still prevalent, and a public health threat to poor rural communities in Malawi despite the ongoing MDA activity. When compared to the findings of other studies done post MDA era, the present findings were slightly higher than was reported in Mangochi (31.5%) in 2020 by Kayuni and friends (4), particularly when considered that the Mangochi study was a serological study and this was microscopy study. It is also higher when compared to a study done by Moyo and friends (5) in the year 2016 in Nkhotakota district (13%). However, other studies done in Zomba in 2014 by Pullanikkatil and friends reported a higher prevalence (49%) this was despite MDA intervention taking place in the area(9). Another similar study done in Chikwawa by Poole and friends in 2014 reported a higher prevalence (49.5%) compared to the current findings (7).Reports from Kaporo Rural Hospital indicated that Mass Drug Administration (MDA), which is done every year in primary schools, did not occur in the space of 2 years prior to the study as the institution prioritized COVID-19 outbreak. The treatment gap that was created could be the reason for this alarming rate of urogenital schistosomiasis reported. In a similar situation in Zanzibar Tanzania, a good progress was reported in a 7 years mass drug administration until a sudden recrudescence in the disease in a just 1 year treatment gap that was created (9). Another treatment gap created in Yamen due to civil war also resulted in sudden rise in schistosomiasis prevalence in the area (21). Secondly, chances of re-infection in the community are very high considering the fact that the activities that lead into dairy water contact mainly farming and fishing are unavoidable at a rural set up. Parents and caregivers take with them their children in farming activities and fishing therefore predisposing to being re-infected despite taking the drug under MDA. Re-infection in schistosomiasis have been reported to be the culprit in the success of MDA as a cohort study analysis done by Marta Till and friends in 2019 (14). In his findings, he reported a high percentage of those that were given drug in cohort study being re-infected.

The association between schistosomiasis and occupation is well documented in literature. A child whose Parent’s occupation is a rice farmer had significantly high risk to contract the disease in the present findings than other occupations, hence rice farming was considered as a risk factor for schistosomiasis in this study. To the best of my knowledge, no study established an association between urogenital schistosomiasis and rice farming in Malawi. However, studies done elsewhere on snails abundance and infection status established that rice fields harbored abundant infected snails (17,18) hence hypothesizing that rice farming could be a risk factor of schistosomiasis transmission. Rice farming in Kaporo-Ngerenge area is the dominant activity done in both rainy and dry seasons. The area being flat and blessed with two big rivers is flood prone hence very high chance of contaminating almost all water bodies. Moreover, during dry season water from these rivers are used in rice cultivation. It is a routine in the area that every child of school going age is involved in rice farming activity hence the high chance of contracting the disease.

Surprisingly. This study showed that children who got a dose of Praziquantel in the year this study was done had a significantly higher risk of getting urogenital schistosomiasis. The findings contradicted several studies done across east Africa (1,9,19,20). This could be attributed to the 2 years treatment gap that occurred prior to the study, which the Kaporo-rural hospital staff attribute to out of stock of the drug. Moreover, the drug is reported to kill only adult worms and have limited ability to enter into blood circulation to kill the juvenile circulating worms and praziquantel has on average a half-life of between 1-2 hours hence high chances that the drug can miss the target (21). Above that, they are also reported cases of praziquantel resistance across Africa (22) and to the best of my knowledge, no such study has been done here in Malawi, leaving chances high that the same can be the case in Malawi for situation of increasing numbers of schistosomiasis cases post MDA are escalating as a study done in Mangochi by Kayuni et al (4) reported high prevalence of both urogenital and intestinal schistosomiasis hence we cannot completely rely on the drug. The high prevalence of urogenital schistosomiasis in those that received praziquantel would be attributed to chances of re-infection. In village life, parents take each and every school age children to assist in rice farms moreover, they is observed behavior in those that received praziquantel that they are free to engage in any risky activities as they have already taken medication thinking the drug prevents them even from contracting the disease. Similar behavior have been reported in a study done on people’s perception on covid-19 vaccination (14).

### Limitations

This study was done after the wave of SARS-CoV-2, therefore there was great resistance from participants to provide urine sample due to misconceptions about COVID-19 issues.

## Data Availability

All data are available and can be provided upon reasonable request.

## Abbreviations

MDA: Mass Drug Administration
SARS-CoV-2: Severe Acute Respiratory Syndrome Corona Virus type 2
COVID-19: Corona Virus Disease of 2019
WHO: World Health Organization

## Acknowledgement

The author wish to acknowledge Master RO Chisale and Rosheen Mthawanji for technical and financial support from proposal development to the publication of this study. Would like also to acknowledge Karonga district hospital officials for authorization of the study, Karonga district hospital laboratory department and Kaporo rural hospital officials for the support in field data collection and laboratory examination.

## Author contributions

Not applicable

## Availability of data and materials

Data collected for this research will be available on a reasonable request after publication in peer-reviewed journals.

## Declarations

### Ethics approval and consent to participate

Following the principle of the Helsinki declaration on ethics for research on human participants Mzuzu University Research Ethics committee (MZUNIREC) with reference number MZUNIREC/DOR/22/21 approved the study. Parents and guardians of the study participants gave their written informed concert.

### Consent for publication

Not applicable

### Author details

Biological sciences department, Faculty of Science, Technology and Innovations, Mzuzu University, P/Bag 201,Luwinga,Mzuzu 2,Malawi

